# Landmark-based analysis of speech differentiates conversational from clear speech in speakers with muscle tension dysphonia

**DOI:** 10.1101/2022.10.20.22281337

**Authors:** Keiko Ishikawa, Mary Pietrowicz, Sara Charney, Diana Orbelo

## Abstract

This study evaluated the feasibility of differentiating conversational and clear speech produced by individuals with muscle tension dysphonia (MTD) using Landmark-Based Analysis of Speech (LMBAS). Thirty-four adult speakers with MTD recorded conversational and clear speech. Of those, 27 speakers were able to produce clear speech. The recordings of these individuals were analyzed with an open-source LMBAS program, SpeechMark®, MATLAB Toolbox ver. 1.1.2.

The mixed-effect logistic regression model indicated that the burst-onset landmark differentiated conversational speech from clear speech. The LMBAS is a potential approach for detecting the difference between conversational and clear speech in individuals with mild to moderate MTD.

## 1. Introduction

Dysphonia (i.e., disordered voice) frequently reduces the speech intelligibility of affected individuals (Ishikawa et al., 2017). Intelligibility is a clinically important outcome measure as it describes one’s ability to be understood by others. Typically measured through the clinician’s auditory-perceptual judgment, this approach faces several challenges associated with human perception, including speaker familiarity, context familiarity, and variability across clinicians. An acoustic-based approach would provide a solution as it is free of these biases. Identifying a biomarker for intelligibility improvement is the first step toward developing such an approach.

Speakers with a healthy voice can enhance intelligibility by intentionally changing how they produce speech. One of the most well-documented intelligibility enhancement techniques is clear speech, which is elicited by instructing speakers to hyperarticulate speech sounds (Ferguson & Kewley-Port, 2007; Moon & Lindblom, 1994; Bradlow et al., 2003; Smiljanic and Bradlow, 2009). Clinically, clear speech is an attractive therapy technique as it exploits the individual’s previous experience in enhancing intelligibility in challenging communication environments. The technique has been incorporated into therapy programs for individuals with dysarthria (Beukelman, Fager, Ullman, Hanson, & Logemann, 2002), hearing loss (Schum, 1997), and more recently, voice disorders (Gartner-Schmidt et al., 2016). Acoustically, clear speech has been shown to elicit temporal and spectral changes in a speech signal, including a decrease in speech rate and increases in vowel duration (Picheny et al., 1986; Ferguson & Kewley-Port, 2002), overall intensity (Picheny et al., 1986), consonant-vowel ratio (Picheny et al., 1986), vowel space (Ferguson & Kewley-Port, 2007), and spectral energy in 1 – 3 kHz (Smiljanic & Gilbert, 2017).

Landmark-based analysis of speech (LMBAS) holds promise for providing a biomarker for clear speech. LMBAS is a knowledge-based approach rooted in the landmark theory of speech production and perception (Stevens, 2002). The theory proposes that articulatory movement elicits abrupt changes in a speech signal, called *landmarks (LMs)*; linguistic information important for speech recognition is contained in the vicinity of the abrupt changes. The algorithm first identifies the moments that qualify as LMs and then classifies them into specific markers based on their acoustic characteristics. The acoustic rules for each LM are shown in Table 1.

**Table 1.** Acoustic rules for each type of landmark. Note that the symbols and mnemonics are not intended to identify underlying articulatory or phonetic events, only to suggest examples: syllabic, voiced frication, etc.

| Symbol | Mnemonic | Rule |
| --- | --- | --- |
| **+g** | Glottal onset | The beginning of sustained vocal fold vibration, i.e., of periodicity or of power and spectral slope similar to that of a nearby segment of sustained periodicity |
| **-g** | Glottal offset | End of sustained vocal fold vibration |
| **+b** | Burst onset | At least 3 of 5 frequency bands show simultaneous power increases of at least 6 dB in both the finely smoothed and the coarsely smoothed contours, in an unvoiced segment (not between +g and the next -g) |
| **-b** | Burst offset | At least 3 of 5 frequency bands show simultaneous power *decreases* of at least 6 dB in both the finely smoothed and the coarsely smoothed contours, in an unvoiced segment |
| **+s** | Syllabic onset | At least 3 of 5 frequency bands show simultaneous power increases of at least 6 dB in both the finely smoothed and the coarsely smoothed contours, in a voiced segment (between +g and the next -g) |
| **-s** | Syllabic offset | At least 3 of 5 frequency bands show simultaneous power *decreases* of at least 6 dB in both the finely smoothed and the coarsely smoothed contours, in a voiced segment |

LMBAS has been applied to describe acoustic differences associated with dysphonia (Ishikawa, Rao, MacAuslan & Boyce, 2020), speech production disorders in children (Atkins, Boyce, MacAuslan, & Silbert, 2019; Liu 2021; Kalita, Mahadeva Prasanna, & Dandapat, 2018), and depression (Huang, Epps, & Joachim, 2019). A few studies have reported the feasibility of detecting the difference between conversational and clear speech with LMBAS in healthy speakers (Boyce, Bradlow, & MacAuslan, 2005; Boyce, Krause, Hamilton, Smiljanic, & Bradrow, 2013).

The findings from the previous acoustic studies of clear speech suggest that burst and syllabic landmarks may be good candidates for biomarkers that differentiate conversational from clear speech. Burst onset and syllabic onset markers may be particularly promising. These landmarks are generated when spectral power simultaneously increases in multiple frequency bands. Picheny et al. (1986) reported the RMS intensity for obstruent sounds, particularly stop consonants, was as much as 10 dB greater in clear speech than in conversational speech. The burst onset landmark is designed to capture such a change in unvoiced regions. On the other hand, the syllabic onset landmark would capture the power increase in voiced regions, as reported by Smiljanic and Gilbert (2017).

The overarching goal of this project is to develop an automatic tool for monitoring patient progress in speech production, which ultimately leads to improved intelligibility. A biomarker incorporating theories of speech production and perception is attractive because it provides better explainability to a machine-learning model than other statistically-derived features. Existing literature suggests the clinical utility of the LMBAS for detecting the difference between conversational and clear speech produced by individuals with dysphonia. Therefore, this study aimed to evaluate whether the LMBAS can differentiate conversational speech from clear speech produced by individuals with muscle tension dysphonia (MTD), a common voice disorder caused by excessive tension in the laryngeal muscles. We tested the following hypothesis: *clear speech will generate a greater number of burst and syllabic onset LMs than conversational speech*.

## 2. Methods

### 2.1. Participants

The participants of this study were thirty-four treatment-seeking adults with MTD who agreed to participate in a randomized clinical trial of voice therapy involving gargle phonation. Of the thirty-four participants, twenty-six were adult females, and eight were adult males. The average age of the participants was 53.5 years old (SD± 18). Fifteen participants were diagnosed with primary MTD (i.e., excessive laryngeal muscle tension as the primary cause of dysphonia), and nineteen participants had secondary MTD (i.e., excessive laryngeal tension that occurs as a result of another underlying laryngeal pathology). The diagnoses were established through clinical voice evaluation by a laryngologist and a speech-language pathologist (SLP). All participants were native speakers of American English and had no history of other speech-language disorders, neurological voice disorders, or hearing loss.

### 2.2. Recording procedure

The participants were seated in a clinical room and fitted with a headset microphone (C555L, AKG). The microphone was placed at 5 cm and 45 degrees off-axis from the corner of the mouth. All speech was recorded directly to a solid-state recorder (TASCAM DR-40X) and digitized at the sampling rate of 44.1 kHz. The SLPs instructed the participants to read the six Consensus Auditory-Perceptual Evaluation of Voice (CAPE-V) sentences, first in the conversational speech style, and then repeat them in the clear speech style. The conversational speech was elicited by asking the participants to “please read these sentences as if you are speaking with a very good friend or family member.” Clear speech style was elicited by asking the participants to “please read these sentences again but this time read them as if you are leaving a very important message on someone’s phone. Please articulate clearly so they can fully understand what you are saying.”

### 2.3. Perceptual analysis for determining dysphonia severity

The severity of dysphonia was rated by four SLPs who specialize in the treatment of voice disorders using CAPE-V. The recorded samples were presented to the SLPs via an online survey created in Qualtrics. The SLPs were instructed to listen to the recordings with headphones in a quiet room. They were allowed to take a break at any time they wished.

### 2.4. Perceptual analysis for speech styles

A listening test was conducted to check the perceptual validity of clear speech. The recordings of conversational and clear speech from the same speaker were presented to two SLPs who specialize in the treatment of voice disorders. The order of the speech style was randomized. The SLPs were asked to identify which recording was more clearly articulated, or whether both recordings had the same level of clarity. The samples that SLPs correctly identified were considered valid productions of conversational or clear speech. The recordings were presented via Qualtrics. The SLPs were instructed to listen to the recordings using headphones in a quiet room and were allowed to listen to them as many times as they wished.

### 2.5. Acoustic analysis

The recordings were manually edited to extract CAPE-V sentences in both speech styles. The starting and ending points of a sentence were visually determined on a spectrogram. Praat (Boersma & Weenink, 2018) was used for the editing process. The listeners in the perceptual analysis described above agreed on 28 out of 32 samples as having a perceivable difference between conversational and clear speech. Only these samples were subjected to the landmark-based analysis using SpeechMark® MATLAB Toolbox ver. 1.1.2. (https://speechmrk.com/).

### 2.6. Statistical analyses

Perceptual data on dysphonia severity ratings: The average was calculated for the ratings from all SLPs. A two-way mixed-effects model intraclass correlations test (ICC) was used to evaluate the inter- and intra-rater reliabilities of the SLPs. Twenty percent of the data were used to evaluate the intra-rater reliability.

Acoustic data: A mixed-effect logistic regression model was used to evaluate the effect of speech style on the landmark count. The speaker participant and sentence were treated as the random effects, and LMs were treated as the fixed effects.

This study was approved by Mayo Clinic’s IRB (Mayo IRB# 20-004267).

## 3. Results

### 3.1. Dysphonia Severity Rating

Table 2 describes the dysphonia severity rating of the participants. All participants’ voices were rated to have a severity below 60 in all rating categories. Within those, 88.24% were rated to have an overall severity of equal to or less than 40 (equal to or less than moderate dysphonia).

**Table 2.** Descriptive statistics of CAPE-V ratings by SLPs. The values indicate the percentage of participants who fell into the specific range of severity.

| Rating category | Rating (0 = normal, 100 = the most severely deviant) |  |  |  |  |  |
| --- | --- | --- | --- | --- | --- | --- |
|  | 0-10 | 11-20 | 21-30 | 31-40 | 41-50 | 51-60 |
| Overall severity | 14.71 | 38.24 | 17.65 | 17.65 | 2.94 | 8.82 |
| Breathiness | 58.82 | 14.71 | 11.76 | 2.94 | 2.94 | 8.82 |
| Roughness | 50.00 | 20.59 | 20.59 | 5.88 | 2.94 | 0.00 |
| Strain | 52.94 | 23.53 | 11.76 | 5.88 | 2.94 | 2.94 |
| Pitch | 91.18 | 0.00 | 5.88 | 2.94 | 0.00 | 0.00 |
| Loudness | 88.24 | 11.76 | 0.00 | 0.00 | 0.00 | 0.00 |

The inter-rater reliability among four SLPs is shown in Table 3. The reliability was highest for overall severity, with an ICC(3,1)=0.74, followed by breathiness, ICC(3,1)=0.70, strain, ICC(3,1)=0.57, pitch, ICC(3,1)=0.56, roughness, ICC(3,1)=0.45, and loudness, ICC(3,1)=0.35.

**Table 3:** Results of the mixed-effect logistic regression model. The asterisk indicates a statistically significant variable.

| Random effects: |  |  |  |  |
| --- | --- | --- | --- | --- |
| Groups | Name | Variance | Standard Deviation. |  |
| Speakers | (Intercept) | 0 | 0 |  |
| Sentence | (Intercept) | 0.1617 | 0.4021 |  |
| Fixed effects: |  |  |  |  |
|  | Estimate | Std. Error | z value | Pr(>\|z\|) |
| (Intercept) | -2.5348 | 0.6455 | -3.927 | 8.61E-05 |
| Total | -0.2278 | 0.1823 | -1.25 | 0.21 |
| [+/- g] | 0.614 | 0.3708 | 1.656 | 0.098 |
| [+b] | 0.4478 | 0.2093 | 2.14 | 0.032* |
| [-b] | 0.2618 | 0.1903 | 1.375 | 0.169 |
| [+s] | 0.4038 | 0.208 | 1.942 | 0.052 |
| [-s] | 0.2906 | 0.2063 | 1.408 | 0.159 |

All raters had high intra-rater reliability ranging from ICC(3,1) of 0.74 to 0.88.

### 3.2. Landmark Analysis

Figure 2 displays the comparison in the average number of LMs generated by conversational and clear speech. On average, conversational and clear speech generated a total of 139.74 (SD±24.48), and 165.67 LMs (SD±26.96) per sentence, respectively. For LM subtypes, conversational speech generated the average of 31.93 [+g/-g] LMs (SD±8.32), 24.96 [+b] LMs (SD±6.03), 20.67 [-b] LMs (SD±6.92),13.07 [+s] LMs (SD±4.79), and 14.41 [-s] LMs (SD±5.34), while clear speech generated the average of 38.33 [+g/-g] LMs (SD±9.33), 30.96 [+b] LMs (SD±7.08), 24.30 [-b] LMs (SD±5.96), 15.14 [+s] LMs (SD±6.27), and 16.00 [-s] LMs (SD±4.73).

**Figure 1:**
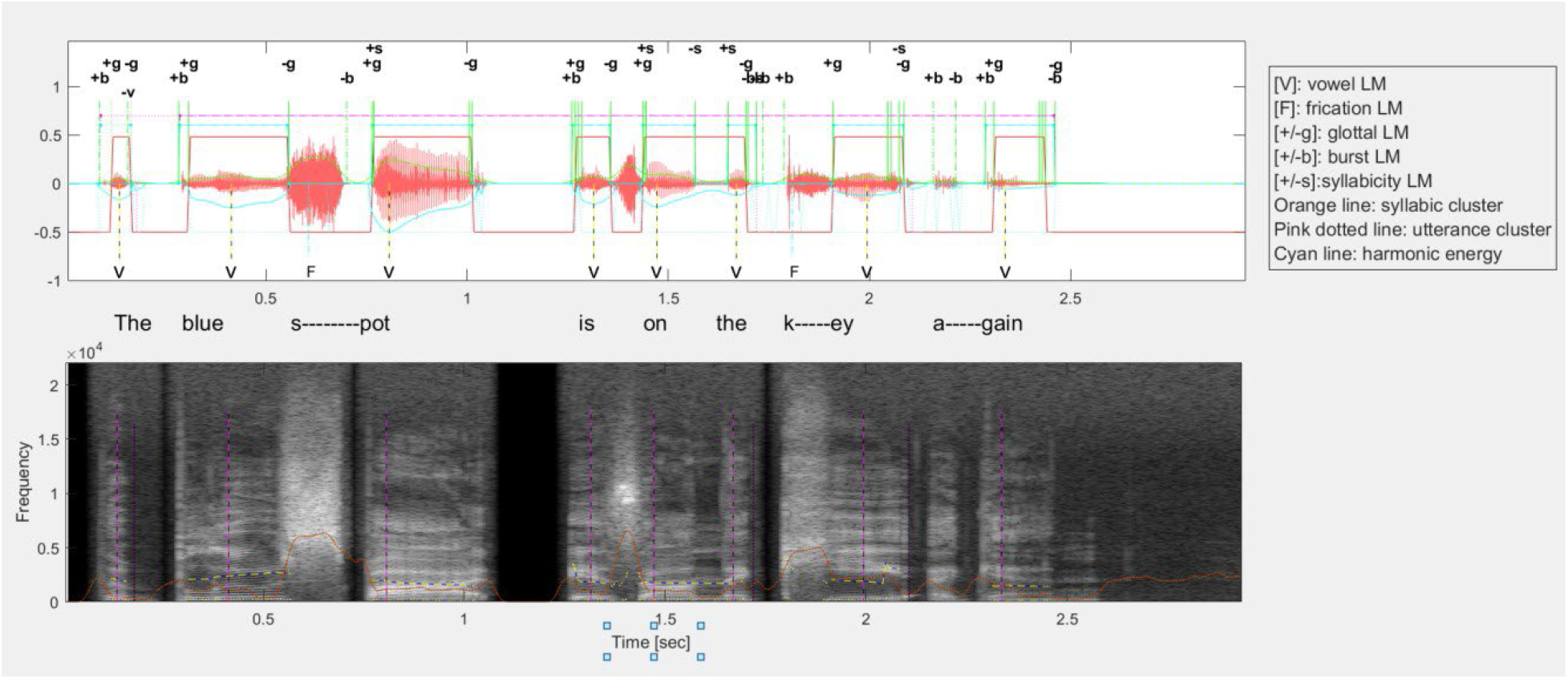
An example of the LMBAS output.

**Figure 2:**
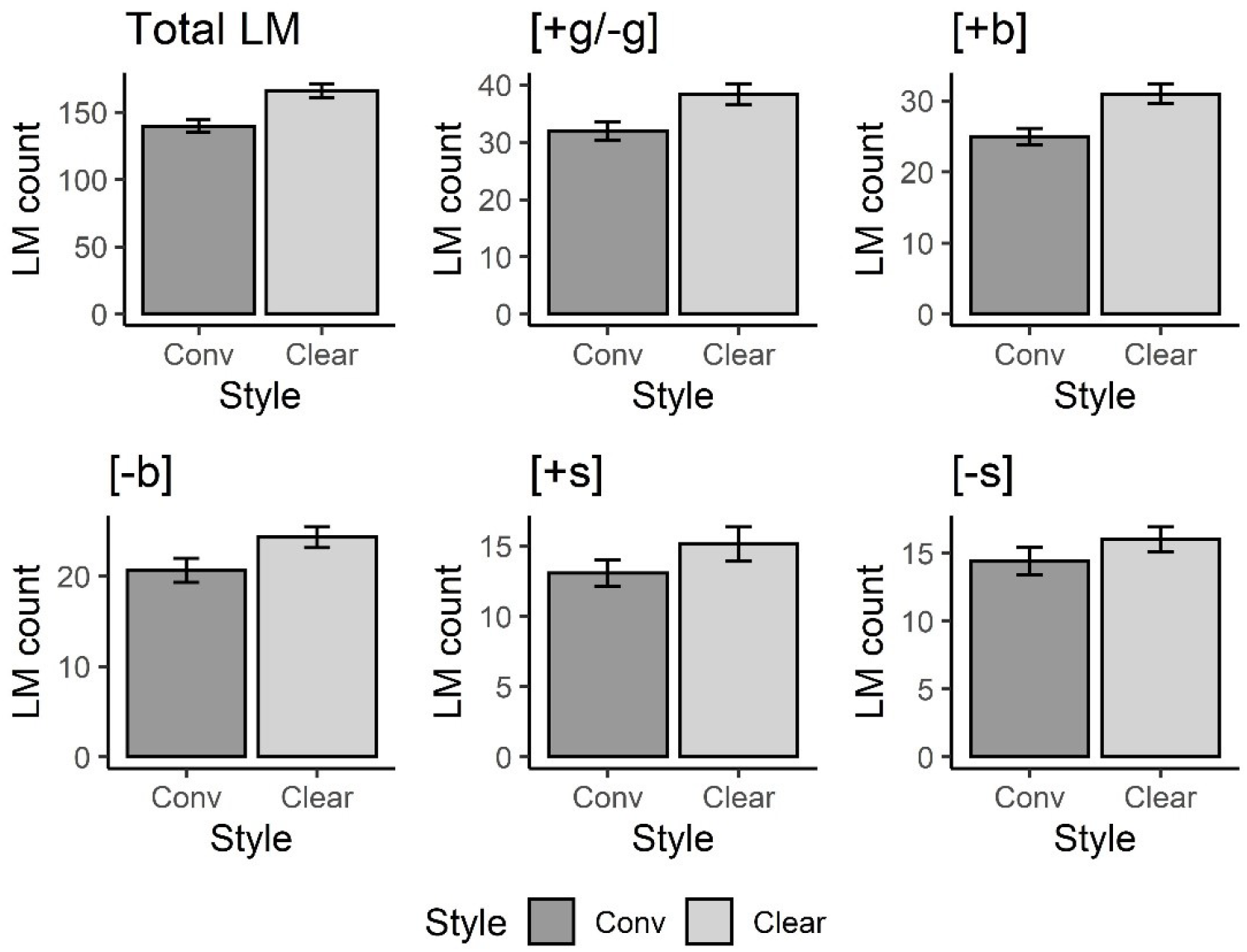
Boxplot showing the average count of LMs. Error bars indicate standard error.

Mixed-effect logistic regression model indicated that clear speech generated significantly more burst onset LMs than conversational speech (Table 3). Since the number of glottal onset and offset landmarks were exactly the same, they were treated as one variable. The threshold of significance was set at *p*=0.05.

## 4. Discussion

This study aimed to evaluate whether the LMBAS would detect acoustic changes associated with clear speech produced by individuals with MTD. Our hypothesis was that the burst and syllabic onset LMs would differentiate conversational from clear speech. The results partially supported the hypothesis: An increase in the number of burst onset landmarks predicted clear speech, while syllabic onset landmarks did not.

This finding may reflect the way our participants implemented clear speech. MTD often reduces the intensity range of one’s voice. The voicing difficulty likely restricted our participants from making an acoustic change required for the generation of syllabic onset LM, which is to increase the spectral energy of their voice across multiple frequency bands. The p-value of this LM was approaching the threshold of significance (*p*=0.052), suggesting that a study with a larger sample may implicate its utility. The statistical significance that the burst onset LM achieved suggests that voicing difficulty likely had less effect on the participant’s ability to emphasize consonants, especially in unvoiced regions. It is also possible that the voicing difficulty made our participants depend more on hyper-articulation of consonants for producing clear speech, resulting in an increase in the burst onset landmark. Despite the general increase in the number of LMs across the LM types, the total number of LMs did not predict clear speech. The reason for this discrepancy is unclear, but possibly due to the small sample size.

Several methodological limitations should be noted. The order of speech style was not randomized during the recording. All participants recorded conversational speech first and then clear speech. The practice effect may have over-inflated the participants’ ability to produce clear speech. Additionally, the dysphonia severity of the participants was limited to mild and moderate. Whether individuals with severe dysphonia would be able to produce clear speech that is noticeable to listeners and whether the burst landmark would continue to predict the speech production change needs to be examined in future studies. Lastly, speech material limits the generalizability of the results in real-world communication scenarios. The recordings used for this study were from sentence reading. Spontaneous speech likely has greater frequency and intensity ranges, which may affect the LMs.

The strength of the LMBAS is that it is a theoretically-driven, knowledge-based system that allows users to evaluate the link between speech production and perception. For future work, the performance of LMBAS can be compared to the performance of other acoustic features commonly extracted for use in voice quality analysis, and other neuropsychiatric disorders that commonly affect voice and emotion recognition (Aguiar et al, 2019; Agurto et al, 2019; Agurto et al, 2020; Bone et al, 2017; Cummins et al, 2015; Deshpande et al, 2020; Eyben et al, 2010; Harati et al, 2018; Huang et al, 2018; Konig et al, 2015; Low et al, 2020; Maor et al, 2020; Marmar et al, 2019; Norel et al, 2018; Orozco-Arroyave et al, 2016; Perez et al, 2018; Pinkas et al, 2020; Rusz et al, 2011; Sara et al, 2020). Some of these features include autocorrelation, zero crossing rate, entropy/entropy ratios across targeted spectral ranges, energy/intensity, Mel/Bark Frequency Cepstral Coefficients (MFCC), linear predictive coefficients (LPC), perceptual linear predictive coefficients (PLP), perceptual linear predictive Cepstral Coefficients (PLP-CC), spectral features, psychoacoustic sharpness, spectral harmonicity, F0, F0 Harmonics ratios, jitter/shimmer, and a variety of statistical and mathematical summary measurements for these frame-level values. These measures should be applied to a control population of healthy voices to explore differences between individuals with MTD and individuals with healthy voices. As the research progresses, the dataset could expand to incorporate other voice disorders and other disorders that impact vocal function so that differential analysis is possible.

## 5. Conclusions

In conclusion, the LMBAS detected the difference between conversational and clear speech in patients with mild to moderate MTD. Future studies should examine whether the current findings will generalize to samples collected from spontaneous speech and to individuals with severe MTD, as well as other types of dysphonia.

## Data Availability

All data produced in the present study are available upon reasonable request to the authors.

## Notes

### Competing Interest Statement

The authors have declared no competing interest.

### Funding Statement

This study did not receive any funding.

### Author Declarations

IRB of Mayo Clinic Alix School of Medicine gave ethical approval for this work.

